# Forecasting Seizure Risk over Days

**DOI:** 10.1101/19008086

**Authors:** Timothée Proix, Wilson Truccolo, Marc G. Leguia, David King-Stephens, Vikram R. Rao, Maxime O. Baud

## Abstract

For persons with epilepsy, much suffering stems from the apparent unpredictability of seizures. Historically, efforts to predict seizures have sought to detect changes in brain activity in the seconds to minutes preceding seizures (pre-ictal period), a timeframe that limits preventative interventions. Recently, converging evidence from studies using chronic intracranial electroencephalography revealed that brain activity in epilepsy has a robust cyclical structure over hours (circadian) and days (multidien). These cycles organize pro-ictal states, hours-to days-long periods of heightened seizure risk, raising the possibility of forecasting seizures over horizons longer than the pre-ictal period. Here, using cEEG from 18 subjects, we developed point-process generalized linear models incorporating cyclical variables at multiple time-scales to show that seizure risk can be forecasted accurately over days in most subjects. Personalized risk-stratification days in advance of seizures is unprecedented and may enable novel preventative strategies.

## Introduction

Despite decades of progress in seizure prediction, reliable methods to mitigate the looming threat of seizures remain elusive (Mormann *et al*., 2007). An ideal prediction model captures all seizures (high sensitivity) while minimizing time in warning (high specificity) (Kuhlmann *et al*., 2018). The pioneering NeuroVista trial demonstrated feasibility of a cEEG-based pre-ictal warning system (Cook *et al*., 2013), but the device used is no longer available. Another implanted device (RNS^®^ System) that uses brain-responsive neurostimulation to treat seizures is available in the U.S. and provides a limited form of cEEG (Geller, 2018). Based on years-long recordings from this device, we recently uncovered daily (circadian) and multi-day (multidien) cycles of IEA that co-modulate with seizure risk (Baud *et al*., 2018). Other factors that putatively influence seizure timing include recent seizure history (Cook *et al*., 2014), sleep duration (Samsonsen *et al*., 2016), and days of the week (Karoly *et al*., 2018). Therefore, we hypothesized that pro-ictal states result from alignment of cyclical influences at ultradian (shorter than a day), circadian, and multidien timescales. To test this, we built PP-GLMs incorporating multiscale cyclical covariates (hereafter “temporal features”) and used an existing dataset to determine whether these models could generate accurate, pseudo-prospective seizure risk forecasts over 24-hour and one-hour horizons (hereafter “daily” and “hourly” forecasts, respectively). PP-GLMs provide a flexible statistical framework to study the association between a sequence of binary events occurring at discrete times (e.g. seizures) and a set of temporal features upon which event probability may depend (Truccolo *et al*., 2004).

## Materials and Methods

### Subjects and data acquisition

We analyzed retrospective cEEG data (227–1049 days) from 18 adults with medically-refractory focal epilepsy who were implanted with the RNS^®^ System (NeuroPace, Inc., Mountain View, CA) at one of two comprehensive epilepsy centers (University of California, San Francisco, N=13 and California Pacific Medical Center, N=5; Supplementary Table 1 and Supplementary Fig. 2). Indications for treatment with the RNS System included bilateral seizure onsets (temporal and frontal), seizures arising from eloquent cortex (visual or motor), and seizure focus contralateral to a prior resection. The study was approved by the Institutional Review Boards at both centers, and all subjects provided written informed consent for participation.

**Figure 1.**
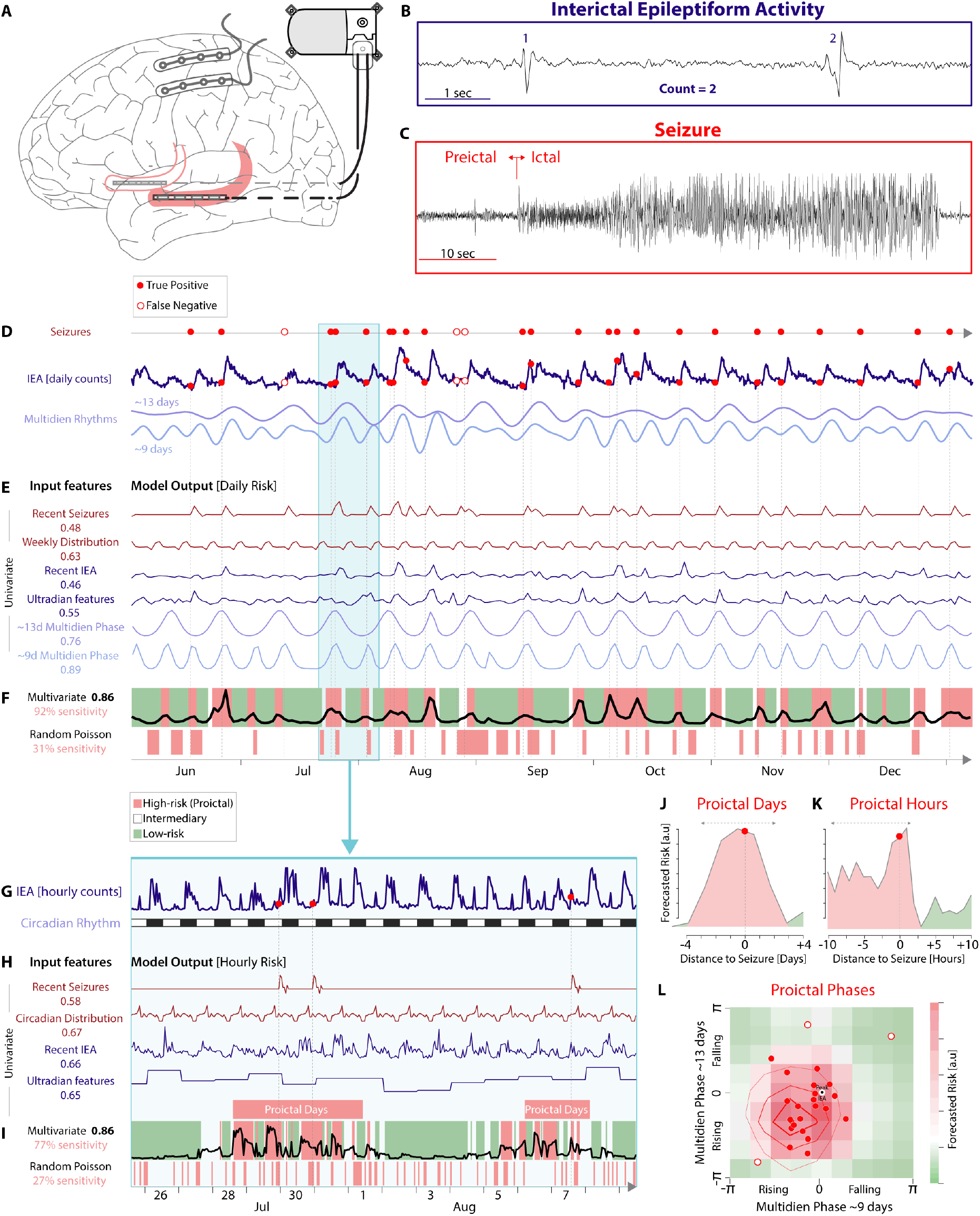
Seizure risk forecasting in one subject (S7). (**A**) RNS System. A cranially-implanted neurostimulator connects to two four-contact intracranial depth leads (bilateral hippocampi in red) and/or cortical strip leads (shown unconnected). (**B**) RNS System cEEG includes hourly counts of IEA detections (here, count=2 for detection of spike-wave discharges) and (**C**) electrographic seizures (note different timescale). ‘Pre-ictal’ refers to the seconds to minutes before seizures. (**D**) Top, seizures timings (red dots) as a point process realizations on a timeline. Middle, seizures in relation to daily IEA. Bottom, multidien cycles derived from IEA time-series with 9- and 13-day periods. (**E**) Univariate daily forecasting using temporal features derived from the seizure (dark red) or IEA time-series (purple-blue) with number corresponding to AUC STiW (see Fig. 2). Model outputs shown over 7 months of held-out test data (training over 10 months not shown). (**F**) Top, multivariate forecasting based on combined temporal features with number corresponding to AUC STiW. Thresholding of model output classifies periods of high (red) and low (green) seizure risk with corresponding sensitivity in red. Most seizures (vertical dotted lines) fall within high-risk periods (filled red dots, true positives) but some do not (empty red dots in D, false negatives) resulting in 92% sensitivity. Bottom, for control, the timing of high-risk periods is treated as realizations of a homogeneous Poisson process (Snyder *et al*., 2008). (**G** to **I**) Similar computations as in D to F, respectively, but using hourly time-series and temporal features derived thereof. (**J** and **K**) Seizure risk forecasts around the time of a seizure (t=0) illustrate pro-ictal periods (red shading) lasting days (J) and hours (K). (**L**) Heat map of forecasted seizure risk as a function of phase combinations of two multidien cycles (D). Contour lines represent actual seizure distribution (red dots as in D), showing that both seizure risk and realization of this risk are maximal when the rising phases of multidien cycles align.

**Figure 2.**
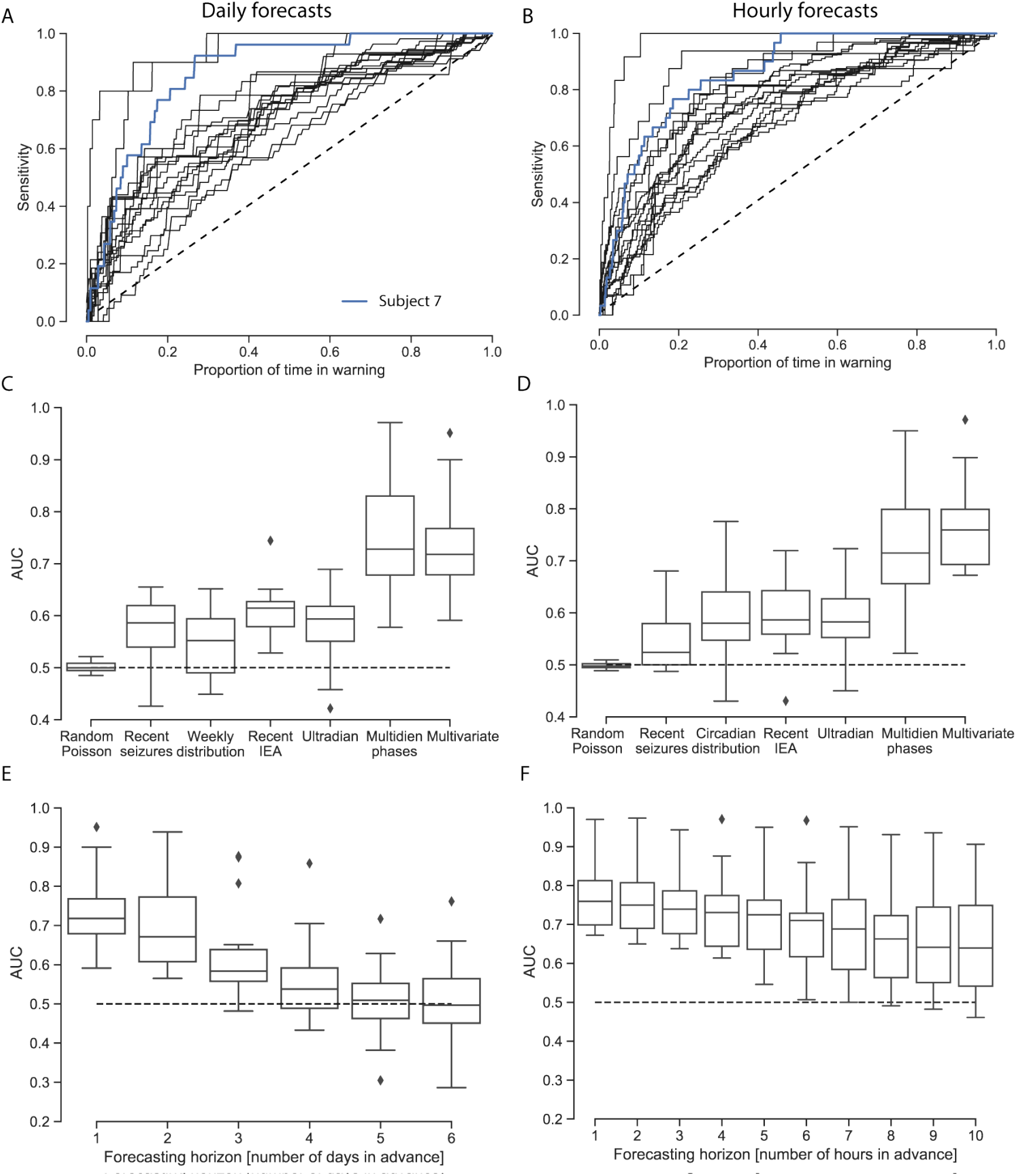
Daily and hourly seizure forecasting performance across all subjects. (**A**) Individual sensitivity vs. proportion of corrected time in warning (STiW) curves showing performance of daily forecasts using combined temporal features on held-out test data (N=18). Blue curve corresponds to the subject shown in Fig. 1. (**C**) Area under the curve (AUC) for STiW when using temporal features alone and combined. (**E**) Model performance as a function of forecasting horizon. (**B, D**, and **F**) Same as A, C, and E, respectively, but for hourly forecasts. Boxplots show median and upper and lower quartiles with whiskers extending to the distribution extremes, outliers (diamonds) excluded. Dotted line shows chance level for a Poisson process.

### Data selection

The primary inclusion criteria for this retrospective study were more than 6 months of available data without large gaps and reliable seizure detection (fig. S2). The RNS System utilizes embedded algorithms based on line-length, area under the curve, and band-pass filters that are iteratively tuned by clinicians to improve sensitivity and specificity of detections of pathological brain activity. For detection of electrographic seizures, we relied on counts of LE, prolonged detections of abnormal activity that often represent seizures. LE timestamps are stored continuously by the neurostimulator even though storage of the corresponding electrocorticograms is limited. As described in detail previously (Baud *et al*., 2018), we only used data from subjects for whom the positive predictive value of LE’s being electrographic seizures was ≥90%. An additional exclusion criteria was >50% of days with seizures, because the utility of forecasting is likely low in individuals with very frequent seizures. For each subject, IEA time-series from two detectors were selected for periods of continuous data with stable detection settings lasting longer than six months. Four detectors can be independently programmed on the neurostimulator, each with a unique parametrization of the embedded algorithm, and Boolean operators (“AND”, “OR”) are used to combine detectors (Sisterson *et al*., 2019). One subject (S15) only had one active detector. For all subjects, the first few months of cEEG (median 150 days) after RNS System implantation were discarded to avoid periods of time when recordings are unstable (Ung *et al*., 2017; Sun *et al*., 2018) and to account for time needed by clinicians to optimize detection parameters (Sun and Morrell, 2014).

### Data pre-processing

Data were pre-processed as described previously (Baud *et al*., 2018). Briefly, changes in detection settings affect detection sensitivity and, therefore, absolute IEA counts. Hourly IEA counts were z-scored by block, where a block corresponds to an epoch with stable detection settings. In six subjects, 8 gaps in the datasets owing to subjects’ non-compliance with data transmission were interpolated (median width 53 hours, range 18–303 hours) as follows: (i) For each gap, we selected flanking data on each side with the same length as the gap. We linearly interpolated the mean value of these windowed data and added a Gaussian random noise with standard deviation (SD) given by the SD of the concatenated IEA data in the two windows; (ii) We used this time-series to compute the different multidien rhythms *x*_multidien,*t*_ using a Morlet wavelet transform; (iii) We computed the circadian distribution of the mean and SD of the IEA time-series and created two corresponding periodic time-series, *x*_meanCircadian,*t*_ and *x*_sdCircadian,*t*_; (iv) Finally, we filled in the gaps by summing the multidien and circadian time-series according to the following equation: *x*_IEAFilled,*t*_ =*x*_multidien,*t*_ + *x*_meanCircadian,*t*_ + *x*_sdCircadian,*t*_ ϵ_*t*_, with *ε*_*t*_ ~ 𝒩 (0,1) the Gaussian distribution. We filled corresponding gaps in the seizure time-series by replacing missing data with zeros.

### Temporal feature extraction

Temporal features were extracted from the interpolated IEA and seizure time-series. The distribution of seizures over 24-h clock time and the calendar week were computed on 60% of the data (training and validation set), and cyclical temporal features were constructed accordingly. We estimated the circadian and multidien IEA cycles using a centered bandpass finite impulse response causal filter of order 2160 and a Hamming window. The bandwidth of the filter was set as *b* = [2*/*3*m*,4*/*3*m*], where *m* is the peak periodicity previously derived by wavelet transform. The cosine and the sine of the phase, as well as the amplitude of each circadian and multidien rhythm, were then extracted through a Hilbert transform of the filtered signal after removal of the mean to compensate for slow drifts. The ultradian features were computed from the IEA time-series using a sliding window of 24 h, recursively shifted by one hour (95.83% overlap). At each time step, if the IEA count was higher than the number of counts averaged over that window, we considered that point as a proxy for sleep. A binary vector representing sleep/non-sleep states was then computed, along with the number of hours of sleep during the preceding day.

### Causality

During temporal feature extraction, we took care to maintain potentially causal temporal relationships so that features at time *t* do not use current or future values of the IEA and seizure time-series. Specifically, we preferred a causal filter to a wavelet transform to estimate the instantaneous phase used in our forecasting model. Ultradian features were computed using only past values of the IEA time-series.

### Point-process Generalized Linear Models (PP-GLMs)

We considered the sequence of seizure occurrence times as a realization of a stochastic discrete-time point process, *S*_*t*_, with the time-bin length set to Δ=1 hour, based on the sampling resolution in our dataset. Because more than one seizure can occur in a given hour (true for 0.2% of the hours on average across subjects), multiple seizure events in a given time-bin were considered as a single seizure event such that *S*_*t*_ ∈ {0,1}. We used PP-GLMs with a log-link function and a (conditionally) Poisson distribution (Truccolo *et al*., 2004) to predict the probability of a seizure (i.e. forecast seizure risk) as a function of features extracted from the most recent seizure history, the most recent history of the IEA, *I*_*t*_, and other covariates {*X*_*t*_ ^1^,*X*_*t*_ ^2^,…}. This probability is related to the “instantaneous” rate or conditional intensity function *λ*(*t*|·) of the point process (Daley and Vere-Jones, 2003), here modeled as:

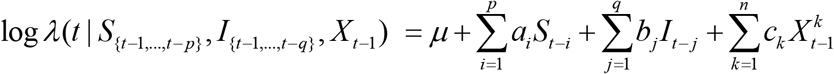

where *p* and *q* correspond to the number of time points for the seizure and IEA histories, respectively; *n* is the number of additional covariates, and *µ* (related to a background rate), *a*_*i*_, *b*_*j*_, and *c*_*k*_ are the model parameters to be estimated. We modelled the conditional intensity as a function of the instantaneous phase *θ*_*t*_ of a multidien cycle as follows:

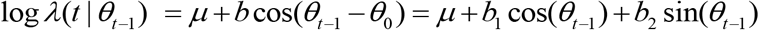

where *θ*_0_ is the preferred phase of the seizure process with respect to the multidien cycle, and *θ*_*t*_ is the corresponding instantaneous phase of the cycle.

Fitting of the model and forecasting was computed with R, using the library tscount (Liboschik *et al*., 2017) (https://cran.r-project.org/package=tscount).

Data were divided into chronological training and held-out test sets comprising 60% and 40% of total data, respectively. To find the optimal length of seizure IEA history for the model, we used five-fold cross-validation: the validation set was chosen sequentially without replacement as 20% blocks of the training data, and the training set consisted of the remaining 80% of data. For models using recent IEA and history, parameter space exploration was run by systematically varying the number of days (0 to 5) and hours (0 to 10) in the history. Optimal parameters were obtained using a performance metric (see below) and used to train PP-GLMs on the whole training dataset. Final performance reported here was assessed on the held-out test dataset.

### Performance metrics

Properly assessing the performance of seizure forecasting models is not a trivial problem (Mormann *et al*., 2007). When classification problems are balanced (e.g. number of seizures equals the number of time points without seizures), model performance is typically evaluated by computing the area under the receiver operating characteristic curve. However, this measure was previously rejected in the field of seizure prediction given that seizures are typically rare events (Snyder *et al*., 2008). For such imbalanced problems, precision-recall curves are more appropriate but tend to underestimate the value of a model when used for seizure forecasting, as they heavily penalize false positives, which could actually represent a true pro-ictal state (e.g. seizure risk being high the day before a seizure). Based on previous developments in the field (Snyder *et al*., 2008), we elected to use a more intuitive measure, the area under the sensitivity (number of seizures correctly predicted divided by total number of seizures) versus proportion of corrected time in warning curve (AUC STiW). The minimum time in warning corresponds to the number of predicted seizures (true positives), as our sampling period (one hour or one day) is equal to our warning duration. As a result, the same model performance would yield lower AUC in subjects with more predicted seizures. We corrected for this bias by using the following definition for the proportion of corrected time in warning:

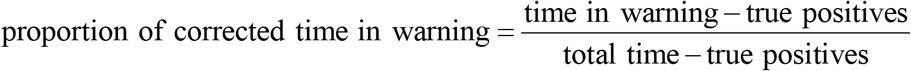

We also report the sensitivity for 25% proportion of corrected time in warning, the proportion of corrected time in warning for 75% sensitivity, and the sensitivity and proportion of corrected time in warning that minimize the Euclidean distance between the curve and the perfect forecasting point (sensitivity = 100% for time in warning = number of seizures).

The proportion of time under low-risk assumed a one-hour or one-day duration and was computed by setting a lower threshold under which only 5% of seizures occurred.

### Statistical analyses

A Poisson process (i.e. memoryless) was used to obtain the sensitivity of a naive, chance-level prediction for the seizure auto-history with a given proportion of corrected time in warning (Snyder *et al*., 2008) (Fig. 1). To determine chance-level forecast of IEA, we used a phase randomization approach to destroy the potential statistical dependence of the seizure point process on this covariate. *M*=200 chance-level surrogate datasets were constructed from the IEA time-series. We used the iterated amplitude-adjusted Fourier transform algorithm (Schreiber and Schmitz, 1996) to randomize the phases of the IEA time-series in Fourier space while conserving the amplitude distribution and the auto-correlation function (power spectrum) of the IEA. Covariates based on each surrogate IEA time-series were then constructed for univariate and multivariate forecasting models. The *p*-values were computed according to *p* = (1+#{*AUC*_*surr*_ *> AUC}*)*/*(1+*M*), where *AUC* was computed on the true test dataset and *AUC*_*surr*_ set was computed based on the chance-level surrogate datasets. We used the false discovery rate to correct for multiple testing (Benjamini and Hochberg, 1995) with a chosen target *α* = 0.05.

## Results

### Multidien rhythms enable seizure forecasting over days

We identified 18 adults (10 males; median age 38, range 20–69) implanted with the RNS System (Fig. 1A) for treatment of mesial temporal (N=13) or neocortical epilepsy (N=5, table S1) and obtained long-term cEEG (median 439 days, range 227–1049 days) comprising hourly counts of IEA and electrographic seizures (Fig. 1, B and C) (Baud *et al*., 2018). We derived temporal features from the IEA time-series, including phase and amplitude of circadian and multidien cycles, and from the seizure time-series, including seizure times over the previous 10 days (auto-history) (Cook *et al*., 2014; Karoly *et al*., 2017) and their circadian and weekly distributions (Griffiths and Fox, 1938; Karoly *et al*., 2018). Our dataset did not include direct sleep measurements (Samsonsen *et al*., 2016), but we used duration of nocturnal IEA increase (an ultradian feature) as a proxy for sleep duration, since IEA typically peaks during normal sleep hours (Spencer *et al*., 2016; Kinnear *et al*., 2018; Frauscher and Gotman, 2019).

PP-GLMs incorporating one (univariate; Fig. 1, D, E, G, and H) or all (multivariate; Fig. 1 F and I) temporal features (see Methods) were trained on 60% (minimum of 17 seizures) of each subject’s hourly or daily (24-hour average) data and tested on held-out data (40% of total, minimum of 12 seizures). Thresholding PP-GLM output probabilities classified periods as low- or high-risk (Fig. 1 F and I), the latter defining hours-to days-long pro-ictal states (Fig. 1, J and K). Even during days of high risk, hours of low risk could be found when taking into account temporal features such as the circadian rhythm (Fig. 1, G to I). PP-GLMs combining phase information from multidien cycles of different period-lengths revealed that seizure risk was highest when the rising phases of these cycles aligned (Fig. 1L).

Performance of the models were quantified using AUC of STiW (Fig. 2, A and B), as described by others (Mormann *et al*., 2007; Snyder *et al*., 2008; Kiral-Kornek *et al*., 2018), and IoC was tested against a model based on a Poisson process (Snyder *et al*., 2008). Across subjects, univariate daily forecasts using temporal features derived from seizure time-series—recent history, circadian time, and weekly distribution—showed modest IoC (Fig. 2, C and D, table S2), likely reflecting the phenomena of seizure clustering (Haut *et al*., 2005), daily peak seizure hours (Langdon-Down and Brain, 1929; Griffiths and Fox, 1938), and preferential days of the week (Karoly *et al*., 2018), respectively. Training models based on recent IEA history, including sleep duration, also produced modest IoC (Fig. 2, C and D). By contrast, the phase of multidien IEA cycles, but not the amplitude, was the dominant contributor to final multivariate hourly and daily models. These multivariate models performed significantly better than surrogate controls (see Methods) for daily and hourly forecasts in 15 (mean AUC=0.73) and 16 out of 18 subjects (mean AUC=0.77), respectively (table S3). Based on our hypothesis of days-long pro-ictal periods, corrected time in warning necessarily lasts several days for each seizure, and AUCs ≥ 0.70 (12 subjects) should be considered as high performance (fig. S1). Three and five subjects had lower but still significant performance for daily (AUC 0.60–0.68) and hourly (AUC 0.65–0.68) forecasts, respectively. The three subjects with lack of IoC had a weak multidien rhythm (S3), a high number of days with seizures (S4, 48%; cut-off was 50%), and relatively short duration of cEEG data (S9, 7.4 months; cut-off was 6 months).

Next, we quantified model performance while increasing the forecasting horizon. IoC was maintained up to three days in advance for daily forecasts (p=1.8 x 10^−6^, two-sided Mann-Whitney U-test) and up to 10 hours in advance for hourly forecasts (p=3.5 x 10^−4^, two-sided Mann-Whitney U-test; Fig. 2, E and F).

### Characteristics of seizure forecasting

To help determine the feasibility of our forecasting approach for future prospective trials, we assessed training requirements for the models, first, by systematically increasing training period length from 12 days to 8 months. Forecasting performance reached 95% of maximum after an average of 101 and 111 days for daily and hourly forecasts, respectively (Fig. 3, A and B). Next, the quality of cEEG signals can fluctuate over time (Ung *et al*., 2017) causing feature drift that may require model retraining (Cook *et al*., 2013). However, performance of our models was stable over a wide range of retraining frequencies (Fig. 3, C and D). Finally, phase-locking values (PLV) between seizures and multidien IEA cycles (Baud *et al*., 2018) moderately correlated with performance of the multivariate forecasting model (Fig. 3, E and F, r=0.56 for days and r=0.63 for hours, Wald test, *p*=0.015 and *p*=0.005, respectively), indicating that retrospective data can be used to estimate forecasting potential for individual subjects.

**Figure 3.**
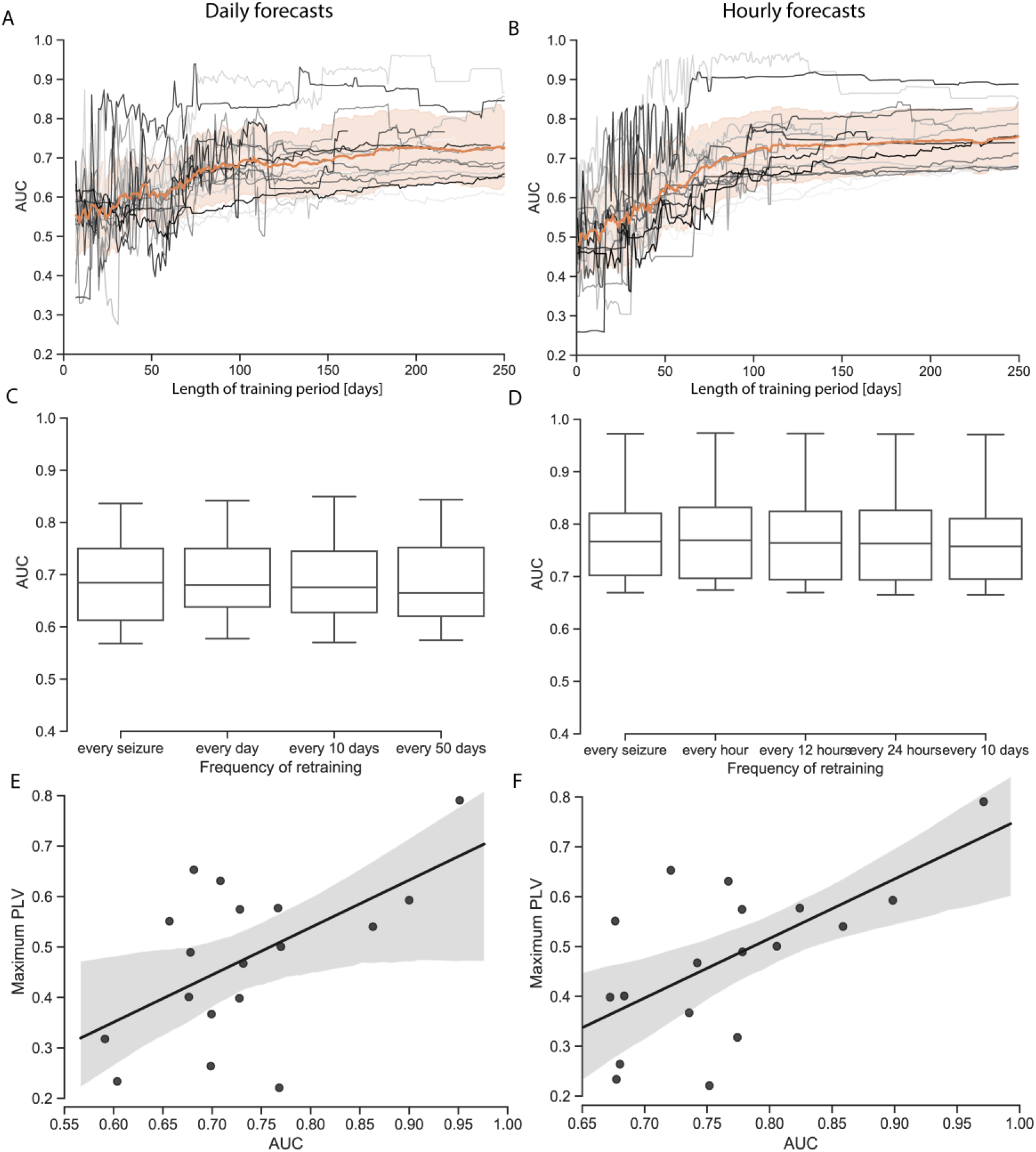
Training characteristics of seizure forecasting models. (**A**) Performance of repeated forecasts on held-out test data using training periods of increasing length. Curves correspond to individual subjects (N=18). Average across subjects in orange, shading ±1 SD. On average, performance plateaus after ∼100 days of training. (**C**) Forecasting performance (AUC STiW) with different intervals of retraining. (**B, D**) Same as A, C, respectively, but for hourly forecasts. (**E, F**) Relationship between multidien phase locking value (PLV) and AUC STiW for all subjects’ daily (E) and hourly (F) forecasts. Pearson correlation coefficients *ρ*=0.56 and 0.63, respectively. Higher PLV predicts better model performance.

## Discussion

In summary, we demonstrate for the first time that seizure risk can be accurately forecasted days in advance. The phase of multidien IEA cycles emerged as the dominant temporal feature for daily and hourly forecasts, supporting our hypothesis of days-long pro-ictal states. The multivariate statistical models used to generate seizure forecasts are intuitive, robust, and modest in their training requirements. Forecasting horizons could be extended for days while maintaining IoC. Based on these results, we propose a nested approach to seizure forecasting: (1) Pro-ictal periods are identified 24 h or more in advance based primarily on the phase of multidien rhythms; (2) During high-risk days, hours of highest seizure risk are determined based on circadian seizure distribution, recent seizure history, sleep patterns, and possibly other factors; and (3) Warnings of impending seizures are provided minutes in advance based on real-time detection of pre-ictal EEG features, taking into account prior seizure probability (Cook *et al*., 2013; Karoly *et al*., 2017). Of practical importance is the fact that (1) and (2) are achievable with currently-approved neurotechnologies, low computational power, and do not depend on real-time responses to features detected in the EEG (Stacey, 2018). Ultimately, forecasting risk and warning of impending seizures would be complementary information provided to patients.

To date, there has been only one prospective trial of a fully functional seizure advisory system (Cook *et al*., 2013). Models used in this trial showed IoC for nine out of 15 (60%) subjects. Across the subset of 10 subjects included in the advisory phase, median sensitivity was 60% and median time in warning was 28%. By comparison, our daily forecasts performed better than chance in 15 out of 18 (83%) subjects, with median sensitivity of 68% and median corrected time in warning of 26% (table S3). A key distinction of our work is that the amount of time in warning is aggregated over days-long periods and is to be understood as a risk, not a deterministic state. In contrast to evanescent minutes-long alerts based on real-time detection of pre-ictal periods, our models detect pro-ictal states and provide smoother forecasting profiles while retaining high performance in most subjects (fig. S1). Metaphorically, previous seizure warning systems worked as ground-based lightning detectors that detect electrical tension proximal to imminent discharges, whereas we used a weather satellite approach that tracks trends at multiple scales to determine local risk of a thunderstorm over upcoming days. Sharp warnings are only desirable provided a high degree of confidence, as too frequent transitions may add stress or cause warning fatigue. In a Bayesian framework, prior knowledge of seizure probability (as done here) can boost reliability of seizure warning systems.

Our results have important theoretical implications: neural dynamics leading to seizures may not operate in a regime of self-organized criticality, as previously hypothesized (Osorio *et al*., 2010). Rather, these dynamics are modulated by robust, multiscale rhythms that drive abnormal networks close to a tipping-point or bifurcation, which, when crossed, results in a seizure (Jirsa *et al*., 2014, p. 20). These critical transitions are typically associated with precursor signatures (Scheffer *et al*., 2009; Kramer *et al*., 2012; Jiruska *et al*., 2013; Rings *et al*., 2019), such as critical slowing (Chang *et al*., 2018), and their direct measurement could boost predictability. Additionally, we speculate that multidien fluctuations in measurable neuromodulators (Baud and Rao, 2018) may influence cortical excitability and cause periodic instability.

Our study has limitations. First, the sample size was relatively small and our subjects may not be representative of all persons with epilepsy. Specifically, these subjects received neurostimulation for treatment of seizures, but we previously showed that the cycles modelled here are independent of this stimulation (Baud *et al*., 2018). Second, the study was pseudo-prospective, so conclusions should be regarded as hypothesis-generating rather than clinical evidence (Kuhlmann *et al*., 2018). Still, the fact that our approach relies on an approved device currently implanted in over 2500 patients with intractable epilepsy sets the stage for larger, prospective trials in the future. Third, we did not assess sleep duration in our subjects, and the proxy biomarker we used (nocturnal IEA increase) is likely imprecise. Finally, our models did not incorporate common seizure triggers, such as stress, medication non-compliance, and alcohol, and some false negatives could relate to such precipitating factors (Bartolini and Sander, 2019).

Identifying pro-ictal states days in advance marks a paradigm shift in epilepsy (Kuhlmann *et al*., 2018). Long-horizon forecasts will likely have practical advantages over last-minute warnings, though clinical utility will need to be tested directly in trials. Future work will also involve optimization of forecasting models, integration with multimodal physiological data (Dumanis *et al*., 2017; Vieluf *et al*., 2019), and development of next-generation seizure forecasting systems (Kremen *et al*., 2018; Stacey, 2018) involving less-invasive devices (Weisdorf *et al*., 2018).

## Data Availability

Data analyzed and code created and used in this study will be made available upon request to the corresponding author after final publication of the manuscript.

## Data availability

Data analyzed and code created and used in this study will be made available upon request to the corresponding author after final publication of the manuscript. Figures 2 and 3 were created on the entire dataset comprising 18 subjects. Figure 1 represent one of these 18 subjects.

## Acknowledgments

Part of this research was conducted using computational resources and services at the Center for Computation and Visualization, Brown University.

## Author contributions

M.O.B., T.P., V.R.R. and W.T. designed the study. V.R.R. collected the data. V.R.R. and M.O.B. selected the data. T.P., M.G.L. and M.O.B. performed the analysis under W.T. supervision. T.P., M.O.B., and V.R.R. wrote the manuscript, which all authors edited.

## Abbreviations

AUC: area under the curve
cEEG: chronic intracranial electroencephalography
IEA: interictal epileptiform activity
IoC: improvement over chance
LE: long episodes
PP-GLMs: point-process generalized linear models
STiW: sensitivity vs. corrected proportion of time in warning

## References

Bartolini E, Sander JW. Dealing with the storm: An overview of seizure precipitants and spontaneous seizure worsening in drug-resistant epilepsy. Epilepsy Behav 2019; 97: 212–218.

Baud MO, Kleen JK, Mirro EA, Andrechak JC, King-Stephens D, Chang EF, et al. Multi-day rhythms modulate seizure risk in epilepsy [Internet]. Nat Commun 2018; 9[cited 2018 May 25] Available from: http://www.nature.com/articles/s41467-017-02577-y

Baud MO, Rao VR. Gauging seizure risk. Neurology 2018; 91: 967–973.

Benjamini Y, Hochberg Y. Controlling the false discovery rate: a practical and powerful approach to multiple testing. J R Stat Soc 1995; 57: 289–300.

Chang W-C, Kudlacek J, Hlinka J, Chvojka J, Hadrava M, Kumpost V, et al. Loss of neuronal network resilience precedes seizures and determines the ictogenic nature of interictal synaptic perturbations. Nat Neurosci 2018; 21: 1742–1752.

Cook MJ, O’Brien TJ, Berkovic SF, Murphy M, Morokoff A, Fabinyi G, et al. Prediction of seizure likelihood with a long-term, implanted seizure advisory system in patients with drug-resistant epilepsy: a first-in-man study. Lancet Neurol 2013; 12: 563–571.

Cook MJ, Varsavsky A, Himes D, Leyde K, Berkovic SF, Oâ€^™^Brien T, et al. The Dynamics of the Epileptic Brain Reveal Long-Memory Processes [Internet]. Front Neurol 2014; 5[cited 2019 Jun 23] Available from: http://journal.frontiersin.org/article/10.3389/fneur.2014.00217/abstract

Daley DJ, Vere-Jones D. An introduction to the theory of point processes. 2nd edition. NewYork: Springer; 2003

Dumanis SB, French JA, Bernard C, Worrell GA, Fureman BE. Seizure Forecasting from Idea to Reality. Outcomes of the My Seizure Gauge Epilepsy Innovation Institute Workshop. eneuro 2017; 4: ENEURO.0349-17.2017.

Frauscher B, Gotman J. Sleep, oscillations, interictal discharges, and seizures in human focal epilepsy. Neurobiol Dis 2019; 127: 545–553.

Geller EB. Responsive neurostimulation: Review of clinical trials and insights into focal epilepsy. Epilepsy Behav 2018; 88: 11–20.

Griffiths G, Fox T. Rhythm in epilepsy. The Lancet 1938; 232: 409–416.

Haut SR, Lipton RB, LeValley AJ, Hall CB, Shinnar S. Identifying seizure clusters in patients with epilepsy. Neurology 2005; 65: 1313–1315.

Jirsa VK, Stacey WC, Quilichini PP, Ivanov AI, Bernard C. On the nature of seizure dynamics. Brain 2014; 137: 2210–2230.

Jiruska P, de Curtis M, Jefferys JGR, Schevon CA, Schiff SJ, Schindler K. Synchronization and desynchronization in epilepsy: controversies and hypotheses: Synchronization in epilepsy. J Physiol 2013; 591: 787–797.

Karoly PJ, Goldenholz DM, Freestone DR, Moss RE, Grayden DB, Theodore WH, et al. Circadian and circaseptan rhythms in human epilepsy: a retrospective cohort study. Lancet Neurol 2018; 17: 977–985.

Karoly PJ, Ung H, Grayden DB, Kuhlmann L, Leyde K, Cook MJ, et al. The circadian profile of epilepsy improves seizure forecasting. Brain 2017; 140: 2169–2182.

Kinnear KM, Warner NM, Gersappe A, Doherty MJ. Pilot data on responsive epilepsy neurostimulation, measures of sleep apnea and continuous glucose measurements. Epilepsy Behav Case Rep 2018; 9: 33–36.

Kiral-Kornek I, Roy S, Nurse E, Mashford B, Karoly P, Carroll T, et al. Epileptic Seizure Prediction Using Big Data and Deep Learning: Toward a Mobile System. EBioMedicine 2018; 27: 103–111.

Kramer MA, Truccolo W, Eden UT, Lepage KQ, Hochberg LR, Eskandar EN, et al. Human seizures self-terminate across spatial scales via a critical transition. Proc Natl Acad Sci 2012; 109: 21116–21121.

Kremen V, Brinkmann BH, Kim I, Guragain H, Nasseri M, Magee AL, et al. Integrating Brain Implants With Local and Distributed Computing Devices: A Next Generation Epilepsy Management System. IEEE J Transl Eng Health Med 2018; 6: 1–12.

Kuhlmann L, Lehnertz K, Richardson MP, Schelter B, Zaveri HP. Seizure prediction — ready for a new era. Nat Rev Neurol 2018; 14: 618–630.

Langdon-Down M, Brain R. Time of day in relation to convulsions in epilepsy. The Lancet 1929; 213: 1029–1032.

Liboschik T, Fokianos K, Fried R. tscount : An R Package for Analysis of Count Time Series Following Generalized Linear Models [Internet]. J Stat Softw 2017; 82[cited 2018 Jan 19] Available from: http://www.jstatsoft.org/v82/i05/

Mormann F, Andrzejak RG, Elger CE, Lehnertz K. Seizure prediction: the long and winding road. Brain 2007; 130: 314–333.

Osorio I, Frei MG, Sornette D, Milton J, Lai Y-C. Epileptic seizures: Quakes of the brain? [Internet]. Phys Rev E 2010; 82[cited 2016 Aug 18] Available from: http://link.aps.org/doi/10.1103/PhysRevE.82.021919

Rings T, Mazarei M, Akhshi A, Geier C, Tabar MRR, Lehnertz K. Traceability and dynamical resistance of precursor of extreme events [Internet]. Sci Rep 2019; 9[cited 2019 Jul 12] Available from: http://www.nature.com/articles/s41598-018-38372-y

Samsonsen C, Sand T, Bråthen G, Helde G, Brodtkorb E. The impact of sleep loss on the facilitation of seizures: A prospective case-crossover study. Epilepsy Res 2016; 127: 260–266.

Scheffer M, Bascompte J, Brock WA, Brovkin V, Carpenter SR, Dakos V, et al. Early-warning signals for critical transitions. Nature 2009; 461: 53–59.

Schreiber T, Schmitz A. Improved Surrogate Data for Nonlinearity Tests. Phys Rev Lett 1996; 77: 635–638.

Sisterson ND, Wozny TA, Kokkinos V, Constantino A, Richardson RM. Closed-Loop Brain Stimulation for Drug-Resistant Epilepsy: Towards an Evidence-Based Approach to Personalized Medicine. Neurotherapeutics 2019; 16: 119–127.

Snyder DE, Echauz J, Grimes DB, Litt B. The statistics of a practical seizure warning system. J Neural Eng 2008; 5: 392–401.

Spencer DC, Sun FT, Brown SN, Jobst BC, Fountain NB, Wong VSS, et al. Circadian and ultradian patterns of epileptiform discharges differ by seizure-onset location during long-term ambulatory intracranial monitoring. Epilepsia 2016; 57: 1495–1502.

Stacey WC. Seizure Prediction Is Possible–Now Let’s Make It Practical. EBioMedicine 2018; 27: 3–4.

Sun FT, Arcot Desai S, Tcheng TK, Morrell MJ. Changes in the electrocorticogram after implantation of intracranial electrodes in humans: The implant effect. Clin Neurophysiol 2018; 129: 676–686.

Sun FT, Morrell MJ. The RNS System: responsive cortical stimulation for the treatment of refractory partial epilepsy. Expert Rev Med Devices 2014; 11: 563–572.

Truccolo W, Eden UT, Fellows MR, Donoghue JP, Brown EN. A Point Process Framework for Relating Neural Spiking Activity to Spiking History, Neural Ensemble, and Extrinsic Covariate Effects. J Neurophysiol 2004; 93: 1074–1089.

Ung H, Baldassano SN, Bink H, Krieger AM, Williams S, Vitale F, et al. Intracranial EEG fluctuates over months after implanting electrodes in human brain. J Neural Eng 2017; 14: 056011.

Vieluf S, El Atrache R, Hammond S, Touserkani FM, Loddenkemper T, Reinsberger C. Peripheral multimodal monitoring of ANS changes related to epilepsy. Epilepsy Behav 2019; 96: 69–79.

Weisdorf S, Gangstad SW, Duun-Henriksen J, Mosholt KSS, Kjær TW. High similarity between EEG from subcutaneous and proximate scalp electrodes in patients with temporal lobe epilepsy. J Neurophysiol 2018; 120: 1451–1460.

